# Mapping the socio-technical causes of overheating risk and unequal thermal burden in hospitals in England: a qualitative systems inquiry using AcciMap

**DOI:** 10.64898/2026.09.10.26362630

**Authors:** Davide Filingeri, Hannah Blount, Nuno Koch Esteves, Thomas Daniels, Carlos Aceves-Gonzalez, Katya Brooks, Katie Jenkins, Patrick James, Tracey Sach, Ana Raquel Nunes, Tom Roberts, Chiara Dall’Ora, Mari Carmen Portillo, Ralph Gordon, Peter R Worsley, Victoria Filingeri

## Abstract

**Background:** Hospital overheating arising from more frequent heatwaves threatens healthcare staff wellbeing, patient safety and service continuity, but is often managed as an estates or seasonal problem. We examined how interacting organisational, infrastructural and experiential conditions sustain overheating vulnerability and unequal thermal burden in hospitals in the south of England.

**Methods:** We conducted a six-phase, multi-source qualitative systems inquiry integrating 129 overheating incident reports from two National Health Service (NHS) hospitals in Hampshire, England, six stakeholder interviews, two focus groups with healthcare staff and patients (n=12), and a national stakeholder refinement focus group (n=4). AI-supported thematic analysis was combined with context-mechanism-outcome synthesis and iterative AcciMap construction, to produce an integrated systems explanation.

**Results:** Twenty themes were consolidated into six interacting explanatory pathways: 1) recognition without durable resilience; 2) hidden visibility; 3) workaround-based resilience; 4) unequal adaptive capacity; 5) underuse of emergency preparedness and business-continuity systems; 6) institutional framing. The analysis indicated that, within the hospital settings investigated, overheating persisted despite recognition because ownership, information, resources and delivery capacity were fragmented across system levels. Frontline workarounds sustained care but could conceal thermal burden and defer structural adaptation. Staff and patients had unequal capacity to regulate exposure because of differences in mobility, autonomy, physiology, clinical dependency and occupational role. Existing preparedness systems offered a route from seasonal reaction to anticipatory action, but overheating was not consistently embedded within them. The AcciMap showed how shared factors connected the six pathways and identified coordinated intervention entry points across monitoring, governance, preparedness, service delivery and infrastructure.

**Conclusions:** Overheating in hospitals in England is best understood as a socio-technical patient-safety and adaptation challenge, not a single-domain estates problem. Our analysis indicates that durable resilience requires environmental intelligence linked to accountable decisions, support for staff and patient adaptation, activation of preparedness systems and long-term infrastructure change.

**Key messages:** What is already known on this topic

• Hospitals in England are increasingly exposed to extreme heat, while ageing infrastructure, limited cooling and operational pressures constrain adaptation.

• Systems approaches can reveal contributory factors across organisational levels but have been used mainly to investigate discrete safety incidents rather than recurrent climate-related vulnerability.

What this study adds

• Six interacting pathways explained why recognised overheating risk can persist within hospitals: fragmented delivery capacity, limited visibility, dependence on workarounds, unequal capacity to adapt, underused preparedness systems and institutional framing.

• Continued service delivery can conceal the burden transferred to staff and patients; apparent operational resilience should not be equated with safe or sustainable heat adaptation.

• Combining thematic analysis, explanatory synthesis and AcciMap modelling connected lived experience and operational evidence to system-level intervention entry points for heat adaptation.

How this study might affect research, practice or policy

• Hospitals in England should connect indoor environmental monitoring with staff, patient and operational consequences; embed heat within emergency preparedness and business continuity; and evaluate who can access protective measures.

• Our AcciMap approach provides an avenue for local remapping and development of mechanism-linked interventions to improve hospitals’ heat resilience.

## Introduction

Heatwaves in the UK and Europe are becoming more frequent, intense and consequential for health.^[1–4]^ The Fourth Independent Assessment of UK Climate Risk identifies intensifying heat as a priority risk and calls for healthcare systems, including hospitals, to remain safe and functional during extreme weather.^[5]^ This requirement is not prospective only: national estates data and evidence submitted to the Environmental Audit Committee describe increasing overheating and limited cooling capacity across the National Health Service (NHS) estate, ^[6–7]^ while hospitals have experienced critical disruption during severe heat.^[8]^

Hospitals in England are particularly exposed because much of the estate was designed for a historic climate and mechanical cooling is largely restricted to operating theatres, intensive care and selected specialist areas.^[5–7]^ Active cooling also carries financial, energy and carbon implications, while alternatives such as shading, passive ventilation, fans and personal cooling remain unevenly evaluated in clinical settings. ^[9]^ Climate-resilient healthcare therefore requires more than a technical decision about air conditioning: it must integrate infrastructure, clinical risk, workforce protection, environmental monitoring, emergency planning and decarbonisation. ^[10,11,16,17]^

The consequences extend beyond thermal comfort. Heat strain can increase fatigue and reduce physical work capacity, ^[12–14]^ and staff fatigue is itself recognised as a contributor to patient-safety risk.^[15]^ Previous qualitative research in English hospitals has documented disrupted work, competing priorities and the limitations of existing heatwave preparedness.^[18]^ National adaptation reports similarly identify gaps in workforce support and uneven implementation of preparedness measures. ^[11,16,17]^ These findings establish important components of vulnerability, but provide less explanation of how infrastructure, governance, information, operational pressure and adaptive behaviour combine to reproduce overheating risk.

Overheating is nevertheless often framed as an estates problem, a seasonal comfort issue or an individual responsibility to hydrate and cope. Such framings can obscure patient autonomy, unequal staff access to protective measures and the organisational work required to maintain service continuity. ^[18,22]^ They can also privilege proximal solutions even though responsibility is distributed across national policy, capital planning, regulation, executive governance, emergency preparedness, operational management, clinical services and the built environment.

Patient-safety science cautions against attributing recurrent adverse conditions in complex healthcare systems to single failures.^[19]^ Socio-technical systems analysis instead examines interactions among people, tasks, technologies, organisational structures and external conditions. ^[21,35]^ AcciMap operationalises this perspective by locating contributory factors across system levels and representing their relationships.^[20,33,36]^ However, mapping alone may show where factors sit without fully explaining how a particular context activates behaviour or reasoning and produces an outcome.

We therefore combined AcciMap with context-mechanism-outcome (CMO) reasoning and resilient-healthcare concepts concerning Work as Imagined and Work as Done ^[23–25,28]^, with the aim of identifying and mapping the socio-technical conditions that generate overheating risk and unequal thermal burden in hospitals in England. We asked how factors across the healthcare system interact to reproduce heat vulnerability, and how the resulting model could support context-sensitive intervention development to improve heat resilience.

## Methods

### Study design and setting

We conducted an iterative, six-phase systems inquiry into two NHS hospital trusts in southern England (i.e. University Hospital Southampton NHS Foundation Trust, UHS; and Hampshire Hospitals NHS Foundation Trust, HHFT), supplemented by national stakeholder input. The phases comprised: (1) retrospective analysis of overheating incidents and local heat plans; (2) stakeholder interviews; (3) staff and patient focus groups; (4) realist-informed synthesis; (5) AcciMap construction; and (6) national stakeholder refinement. Each phase informed subsequent sampling, questioning and analysis rather than operating as an independent study component. **Table 1** makes explicit the contribution of each evidence source.

**Table 1.**
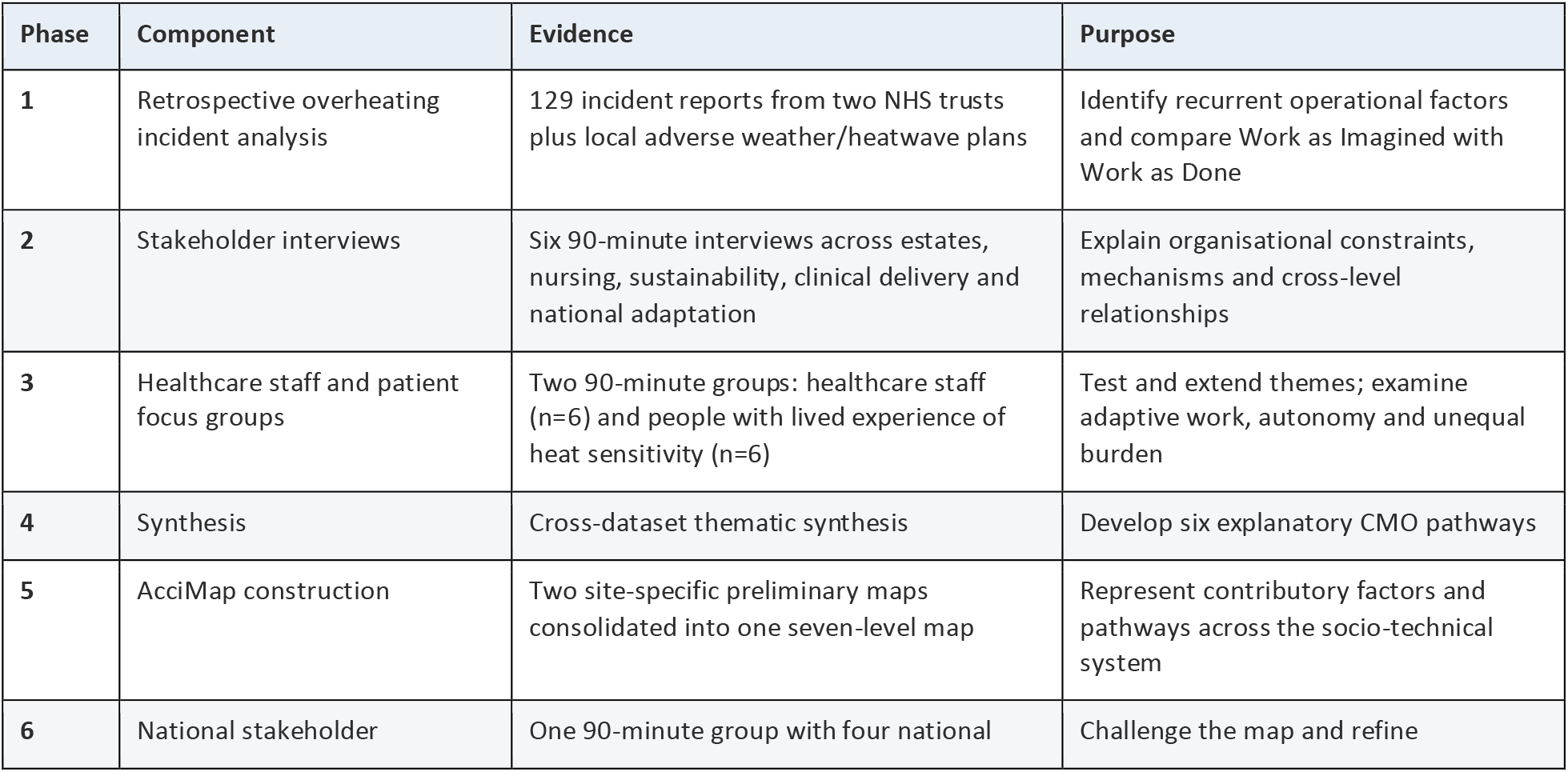
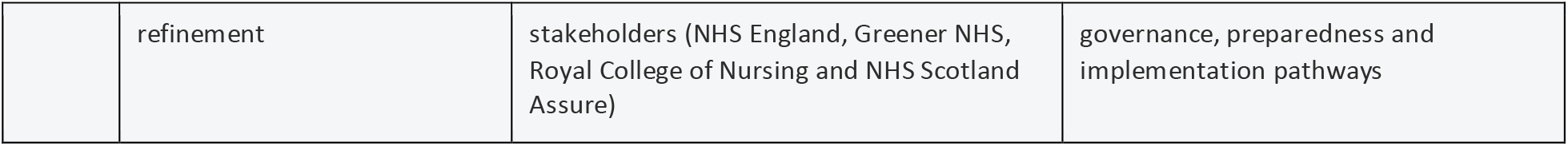
Evidence sources, participant composition and analytical contributions.

The study adopted a critical-realist orientation, treating hospital overheating as an emergent phenomenon produced through interacting material, organisational, cultural and governance conditions. Socio-technical systems theory guided attention across infrastructure, policy, management, workforce practice and patient experience.^[21,35]^ Resilient-healthcare concepts informed interpretation of frontline adjustment and the distinction between formal plans and work as performed. ^[23,25]^ AcciMap provided the systems-modelling structure, ^[20,33]^ and realist-informed CMO reasoning provided the explanatory logic.^[24,28]^ CMO pathways were treated as evidence-supported propositions, not experimentally established causal relations.

The University of Southampton Ethics Committee approved the service evaluation of incident data (ERGOII 111304; also registered with University Hospital Southampton research and development) and the interview and focus-group study (ERGOII 109825). Participants gave informed consent. Transcripts and operational material were anonymised.

### Data sources

We analysed 129 operational reports. UHS supplied 90 estates-helpdesk records relating to overheating during three periods of the 2022 UK heatwave and a comparable 2021 period, together with its Adverse Weather Plan. HHFT supplied 39 incidents recorded under its Severe Weather Events category between April and December 2025, together with its Heatwave Response Plan.^[26,27]^ Records concerned high temperatures, thermal discomfort, ventilation or cooling failure and heat-related operational concerns. The two datasets differed in reporting platform and observation period and were not treated as directly comparable estimates of incidence. Formal plans represented Work as Imagined, while incident narratives provided evidence of Work as Done.

Six 90-minute online semi-structured interviews were purposively sampled to provide maximum variation across estates and facilities, nursing leadership, sustainability, frontline clinical work, combined sustainability/clinical practice and national adaptation planning (**Table 2**).

**Table 2.** Evidence sources and arising perspectives for the 6 semi-structured interviews.

| Perspective | Role | Level/setting |
| --- | --- | --- |
| Estates/facilities | Director of Estates, Facilities and Capital Development | Hospital trust |
| Nursing leadership | Chief Nursing Officer | Hospital trust |
| Sustainability | Head of Sustainability | Hospital trust |
| Clinical delivery | Midwife | Hospital trust |
| Combined operational/clinical role | Sustainability Project Delivery Officer and Recovery Nurse Practitioner | Hospital trust |
| National adaptation planning | Member of Climate Response | Department of Health and Social Care Wales |

Two separate focus groups (n=12) comprised six healthcare staff and six patient/lived -experience contributors. Some contributors also brought former or current healthcare-professional expertise. Detailed participant characteristics are reported in **Table 3**.

**Table 3.** Evidence sources and arising perspectives from the 2 focus groups.

| Group | Participant characteristic/role | Setting or perspective |
| --- | --- | --- |
| Healthcare staff | Matron | Professional |
| Healthcare staff | Physiotherapist | Professional |
| Healthcare staff | Occupational therapist | Professional |
| Healthcare staff | Procurement officer | Professional |
| Healthcare staff | Medical doctor | Professional |
| Healthcare staff | Medical doctor | Professional |
| Lived experience/patient group | Person with spinal cord injury; former physiotherapist | Lived experience |
| Lived experience/patient group | Person with spinal cord injury | Lived experience |
| Lived experience/patient group | Person with spinal cord injury | Lived experience |
| Lived experience/patient group | Person with multiple sclerosis; former physiotherapist | Lived experience/professional |
| Lived experience/patient group | Person with chronic fatigue syndrome; clinical exercise physiologist | Lived experience/professional |
| Lived experience/patient group | Specialty Registrar in Public Health | Lived experience/professional |

A subsequent online focus group purposively sampled four national stakeholders spanning NHS England nursing sustainability, Greener NHS, Royal College of Nursing and NHS Scotland Assure. The final national stakeholder focus group was used primarily to challenge and refine the emerging explanatory model rather than to reopen exploratory theme development. Participants examined omissions, wording, connections, governance feasibility and potential implementation mechanisms. Their contribution particularly refined environmental intelligence, ownership diffusion, preparedness and business-continuity integration, institutional framing and trusted professional communication.

### Analysis and integration

#### Phase 1: operational records and preliminary maps

UHS supplied 90 estates help-desk records covering three periods during the 2022 heatwave and a comparable period in 2021, together with its Adverse Weather Plan. HHFT supplied 39 records from its Severe Weather Events subcategory for April–December 2025 and its Heatwave Response Plan. Both datasets were supplied in December 2025.

Davide Filingeri (DF) and Victoria Filingeri (VF) undertook the initial thematic analysis and developed the codebook. They compared reported events and responses with planned actions, informed by the distinction between Work as Imagined and Work as Done. Records were treated as partial accounts of reported practice, not direct observation or comparable estimates of incident frequency. Differences in periods, reporting systems and categories precluded pooling them as an incidence baseline. Emerging themes and identified factors informed the development of preliminary Trust-level AcciMap(s) and subsequent questioning (**Fig. 1**). Separate published reports describe the operational datasets.^[26,27]^

**Figure 1.**
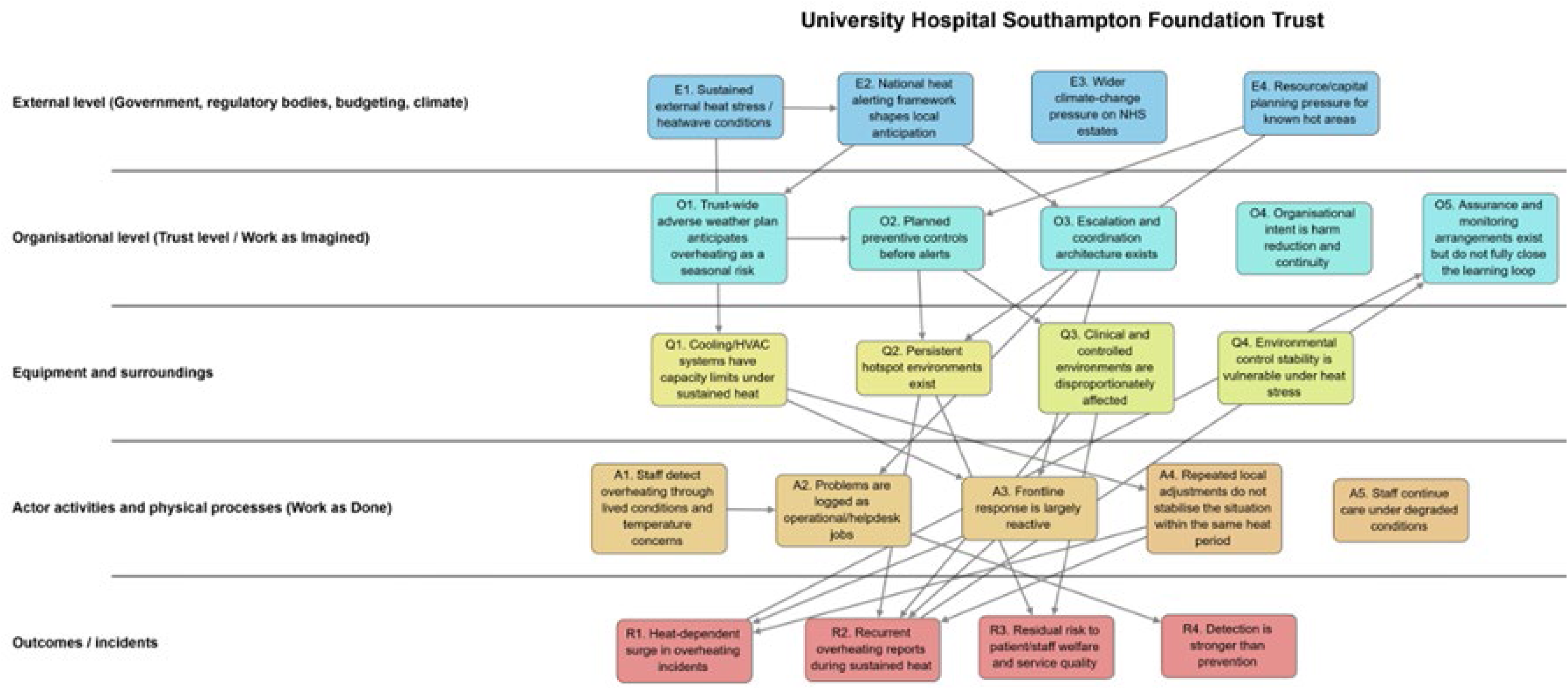
Preliminary AcciMap for University Hospitals Southampton NHS Foundation Trust based on overheating incident data.

#### Phases 2–3: interviews and focus groups

Six semi-structured interviews and two focus groups were conducted online, each lasting approximately 90 minutes and audio-recorded with consent. DF led all the sessions. Interview and focus participants were purposively selected to contribute strategic, operational, frontline, lived experience and policy perspectives. After each session, anonymised transcripts were uploaded to OpenAI GPT-5.6 Sol (San Francisco, California, USA) for thematic analysis using the human-generated codebook. Meaning units comprised one or more sentences conveying a coherent idea. Coding distinguished system location, functional domain and explanatory mechanism, with additional CMO tags. Cross-case matrices supported comparison of recurrent and role-specific patterns, including differences between staff and lived-experience perspectives.

#### Phases 4–5: explanatory synthesis and AcciMap consolidation

OpenAI GPT-5.6 Sol (San Francisco, California, USA) was used to perform an initial cross-dataset synthesis from incident-analysis outputs and transcript analyses. DF and VF reviewed emerging CMO pathways and interpretations against underlying transcripts and discussed their interpretations. Emerging CMO pathways included descriptive organisational processes and candidate explanations; the configurations were used to structure the systems account rather than claim independently established generative mechanisms. DF and VF used the reviewed outputs to revise and consolidate the preliminary Trust-level AcciMap(s). Synthesised themes and pathways informed factor (re-)placement and proposed links within and between system levels. The preliminary AcciMap(s) contained five levels; the consolidated AcciMap distinguished six socio-technical levels—national policy; regulation, oversight and preparedness; executive governance; operational management; staff and patient experience; and infrastructure—with a seventh outcome layer. Shared factors connected pathways where the human-reviewed synthesis indicated interdependence. The final AcciMap integrated recurrent heat vulnerability across events and perspectives rather than reconstructing one adverse event.

#### Phase 6: national stakeholder scrutiny and refinement

DF and VF jointly led a final 90-minute online, recorded focus group with four national stakeholders representing NHS England nursing sustainability, Greener NHS, Royal College of Nursing and NHS Scotland Assure. Participants examined the consolidated AcciMap for coherence, omissions and governance or implementation relationships. DF and VF thematically analysed this dataset using the human-generated codebook. A codebook refinement addendum was developed, documenting retained concepts, revised definitions and implications for the AcciMap. This phase contributed to further interpretation and contextual refinement of the AcciMap. Reinforcement alongside continuing refinement was understood as consolidation, without claiming definitive saturation.

#### Human review and reflexivity

DF and VF met at each phase end to review evidence and interpretations. DF’s direct familiarity with all data collection sessions informed contextual review. Both researchers retained responsibility for the integrated explanation and final AcciMap, attending to assumptions about responsibility, infrastructure and frontline adaptation. Codebook versions, cross-source matrices, CMO tables, maps and the stakeholder addendum documented analytical development. DF and VF also produced a structured quote-extraction matrix. Quotations were selected for mechanism-revealing value, cross-level representation, explanatory strength and contribution to systems interpretation. Selection occurred after cross-dataset synthesis and mechanism stabilisation, providing traceability between empirical material and the final interpretation.

### Patient and public involvement

A project-specific PPIE panel was formed to support all phases of the research underlying this project. The panel comprised eight individuals with lived experience of temperature sensitivity and overheating within hospitals in England and met monthly over the 10-month duration of the project to inform research design, inclusivity, and knowledge mobilization. Individuals with heat-sensitive conditions contributed lived-experience evidence and challenged assumptions about mobility, autonomy and access to adaptation, and informed the interpretation of unequal thermal burden.

## Results

### Integrated themes

Across 129 incident reports, six interviews, two staff and patient/lived -experience focus groups (n=12) and the national refinement group (n=4), overheating emerged as a recurrent socio-technical risk rather than an isolated estates problem. Eight core themes were strongly represented across most or all sources; four strong supporting themes explained institutional and temporal constraints; and eight dataset-specific themes added operational or lived-experience detail (**Table 4**).

**Table 4.** *Complete cross-source thematic evidence matrix, presenting the 20 Integrated themes and evidence source*. ✔ strong evidence; ◑ partial or role-specific evidence; + new contribution or substantive strengthening from a focus group; ✖ not identified. FG: Focus group. Classification reflects breadth and explanatory contribution, not quantitative prevalence.

| No | Integrated theme | Incident data | Interviews | FG1 healthcare staff | FG2 patients/lived experience | Classification |
| --- | --- | --- | --- | --- | --- | --- |
| 1 | Recognition without durable resilience | ✓ | ✓ | ✓ | ① | Core |
| 2 | Infrastructure and estate constraints | ✓ | ✓ | ✓ | ✓ | Core |
| 3 | Recurrent workaround model | ✓ | ✓ | ✓ | ✓ | Core |
| 4 | Data invisibility and under-reporting | ✓ | ✓ | ✓ | ① | Core |
| 5 | Cultural normalisation of heat burden | ① | ✓ | ✓ | ① | Core |
| 6 | Governance awareness but limited implementation capacity | ① | ✓ | ✓ | ✓ | Core |
| 7 | Unequal thermal burden across roles, bodies and conditions | ① | ✓ | ✓ + | ✓ + | Core; strengthened by focus groups |
| 8 | Clinical and safety trade-offs constrain cooling | ✓ | ✓ | ✓ | ✓ | Core |
| 9 | Sustainability/adaptation/safety framing tension | ① | ✓ | ① | ✓ | Strong supporting |
| 10 | Temporal mismatch between immediate pressure and long-term adaptation | ✓ | ✓ | ① | ① | Strong supporting |
| 11 | Participation and inclusion gaps | ① | ✓ | ✓ + | ✓ + | Strong supporting |
| 12 | Tools and guidance exist but need capability and ownership | ① | ✓ | ✓ | ① | Strong supporting |
| 13 | Transient worker exclusion | ✕ | ① | ✓ + | ✕ | Dataset-specific |
| 14 | Dynamic occupancy overload | ① | ① | ✓ + | ✓ | Dataset-specific |
| 15 | Access inequality to wellbeing measures | × | ① | ✓ + | ① | Dataset-specific |
| 16 | Thermal transition stress | × | ① | ✓ + | ✓ + | Dataset-specific |
| 17 | Loss of patient autonomy/inability to self-regulate | × | ① | × | ✓ + | Dataset-specific |
| 18 | Micro-design features shape thermal experience | ① | ① | ✓ | ✓ + | Dataset-specific |
| 19 | Outdoor/restorative spaces as thermal-resilience assets | × | × | ① | ✓ + | Dataset-specific |
| 20 | Patients and staff as co-producers of feasible interventions | × | ✓ | ✓ + | ✓ + | Dataset-specific |

The matrix shows both convergence and analytical extension. Infrastructure and estate constraints, recurrent workarounds and clinical trade-offs were visible in incident records and reinforced by qualitative accounts. Interviews made governance awareness, implementation limitations and institutional framing more explicit. Staff focus groups added transient-worker exclusion, dynamic occupancy, unequal access to wellbeing resources and thermal transitions. Patient/lived -experience evidence added loss of autonomy, condition-specific sensitivity, the importance of air movement and micro-design effects on comfort and dignity. These additions did not sit outside the core analysis: they explained how apparently generic heat exposure became unequally experienced and managed.

The themes were also mutually reinforcing rather than independent categories. Infrastructure constraints increased reliance on workarounds; workarounds sustained care while reducing visible disruption; normalisation and fragmented reporting weakened the organisational signal of burden; and weak visibility reduced the case for investment. Role, mobility, dependency and environmental control determined who could use nominally available adaptations and who absorbed the burden of continuity.

### Six interacting pathways

The themes were synthesised into six CMO-informed pathways, which distinguished identifiable context, proposed explanatory mechanisms and observed or anticipated outcomes (**Table 5**).

**Table 5.** The six pathways and explanatory formulation, their underlying themes from cross-source thematic evidence matrix (i.e. Table 4), and relevant illustrative quotes.

| Pathway and explanatory formulation | Illustrative evidence (quotes) |
| --- | --- |
| **A. Recognition without durable resilience**<br>**Context:** Increasing heat exposure occurs within ageing, climate-inadequate infrastructure and financial, workforce and operational pressures.<br>**Mechanism:** Recognition does not translate reliably into action where adaptation is under-prioritised, ownership fragmented and implementation capacity constrained.<br>**Outcome:** Recurrent vulnerability persists despite awareness and seasonal mitigation.<br>*Underlying themes from cross-source thematic evidence matrix: 1, 2, 6, 10, 12.* | *"So the NHS as an overall organisation has at least recognised that that risk [heat] is coming and will get worse. So I think that's a recognition that we don't want to get to that point where like a catastrophic event is the trigger point for making change"*<br>— Interview, hospital organisational leadership<br>*"Our health and safety report that has gone to board factors in the heat in our ward environment [...] I think we are absolutely aware of the risks. It's not an environment that we'd want people to be working in, but to do something about it, there isn't the money."*<br>— Interview, hospital organisational leadership |
| **B. Hidden visibility**<br>**Context:** Fragmented monitoring and reporting coexist with repeated seasonal overheating and cultures prioritising endurance, continuity and patient care.<br>**Mechanism:** Weak environmental intelligence limits governance visibility; normalisation and routine coping can reduce escalation and reporting signals.<br>**Outcome:** Reduced visibility may weaken prioritisation, investment and coordinated response.<br>*Underlying themes from cross-source thematic evidence matrix: 3, 4, 5.* | *"As a nurse, we kind of just get on with it when it comes to stuff like that. You kind of, oh, well, it's hot. What can we do in the moment?"*<br>— Interview, healthcare staff<br>*"The more you normalise something, the less it's reported. So there will be an element of this is to be expected, this is normal. You know, it's always unbearably hot here"*<br>— Interview, hospital organisational leadership |
| **C. Workaround-based resilience**<br>**Context:** Structural vulnerabilities persist while continuity of healthcare delivery remains essential.<br>**Mechanism:** Staff and operational teams use temporary fixes, reactive cooling and adaptive workarounds to maintain services.<br>**Outcome:** Immediate disruption may be reduced while underlying constraints persist; deferred adaptation is a proposed systems relationship, not an isolated causal effect of coping.<br>*Underlying themes from cross-source thematic evidence matrix: 2, 3, 8, 10.* | *"Generally, it's the same thing each year...we have loads of actions to say, right, we really need to sort all this stuff out because of heat waves. Then we forget about it all until the next year when there's another heat wave"*<br>— Interview, hospital organisational leadership<br>*"Sometimes they come along with fans that you can put on your desk, but because the area is so tight on space already, and then you whack a big fan down, which takes up a load of space on your desk, then you're even more cramped, so it makes things worse"*<br>— Focus group, healthcare staff |
| **D. Unequal adaptive capacity**<br>**Context:** Staff and patients differ in mobility, physiological sensitivity, role flexibility, dependency, environmental control and access to resources; both encounter thermally inconsistent spaces.<br>**Mechanism:** Role constraints and dependency limit self-regulation through clothing, movement, positioning or environmental changes. Thermal transitions were described as an additional burden.<br>**Outcome:** Thermal burden, discomfort, symptoms, cognitive load and loss of dignity are amplified for particular staff and patient groups beyond static room-temperature exposure alone.<br>*Underlying themes from cross-source thematic evidence matrix: 7, 8, 11, 13–19.* | *"When you're like a transient member, [...] you don't have a base, like you're not on just one ward, we cover multiple wards and multiple floors. It's really hard to have your [water] bottle somewhere because you aren't just in one place. I think the opposite is with nurses, it's really hard for them to get off the ward because they're stuck in safe numbers."*<br>— Focus group, healthcare staff<br>*"I think it's only when you do have to step out of the [delivery] room for something, you might then actually appreciate that you are feeling maybe a bit weak or maybe a bit slow in cognitive processing. And although there's not much we can do to cool the [delivery] room necessarily, we also have to be mindful that if the baby is, you know, imminently going to be born or indeed has already been born, we have to keep the room really warm for them."*<br>— Focus group, healthcare staff<br>*"Being too hot or cold is not so much of a problem if you can move."*<br>— Focus group, patient / lived experience |
| **E. Emergency preparedness and business continuity underuse**<br>**Context:** Preparedness structures exist, but adaptation responsibilities span estates, sustainability, workforce, governance and clinical services amid competing priorities.<br>**Mechanism:** Ownership diffusion and unclear accountability can weaken implementation. Embedding heat within business-continuity and preparedness arrangements was proposed as a means of coordinating anticipatory action.<br>**Outcome:** Fragmented or reactive adaptation may persist. Improved coordination and scalability are candidate benefits requiring evaluation.<br>*Underlying themes from cross-source thematic evidence matrix: 6, 9, 10, 12; + national stakeholder refinement.* | *"The other piece of language that I think is quite useful is when we talk about climate adaptation, it's that everybody is pointing at whose job it is. [...] so someone has to hold the ring on this work, but it has to be a multidisciplinary team approach."*<br>— Focus group, national stakeholder<br>*"One of the biggest risks is that consistent reaction, instead of the proactive nature of what we need to do. [...] It's often quite sort of, there is the planning, obviously, but it's to react as opposed to trying to look ahead and think we know this is coming, let's do something. [...] The business continuity process is a way of being able to get ahead of this by linking it to health alerts."*<br>— Focus group, national stakeholder |
| **F. Institutional framing**<br>**Context:** Organisational priorities are shaped by familiar operational language; clinicians and affected staff and patients offer professional credibility and contextual knowledge.<br>**Mechanism:** Framing heat through seasonal pressures, safety and continuity, using trusted clinical voices, and involving users in design were proposed to improve legitimacy and feasibility.<br>**Outcome:** Greater engagement, acceptance and effective adaptation are anticipated possibilities.<br>*Underlying themes from cross-source thematic evidence matrix: 9, 11, 12, 20; + national stakeholder refinement.* | *"I think the original green plan guidance from Greener NHS [...] is brilliant, but it's a 90% focus on emissions reduction with a cursory one section on climate adaptation, and I think the challenge of that is [adaptation] it becomes the poor relative."*<br>— Focus group, national stakeholder<br>*"I would push for seasonal pressures instead of winter or summer [...] actually every season there are pressures that we need to be aware of"*<br>— Focus group, national stakeholder<br>*"But there is something here about utilising the power of the trusted voices of clinicians in the conversations that we're having with patients, whether that's within a trust or a primary care centre or in the community, to be able to engage as the carrier of that message around overheating and then what to do about it in order to support communities"*<br>— Focus group, national stakeholder |

Recognition without durable resilience (pathway A) was illustrated by organisational accounts that combined acknowledged heat risk with limited resources to act. This supports a distinction between recognition and implementation capacity. Hidden visibility (pathway B) concerned normalisation and reduced reporting: staff described getting on with work, while leadership accounts explicitly linked repeated exposure with diminished escalation. The inference is that formal reporting captures only part of the burden, not that the operational records are uninformative. Workaround-based resilience (pathway C) connected temporary mitigation with recurrent unresolved problems. An organisational account described action recurring each summer and then losing momentum; a staff account described how a fan could worsen already constrained workspace. Together, these illustrate the persistence and practical trade-offs of coping. They support the proposed relationship between continuity and deferred adaptation, but do not isolate workarounds as the cause of underinvestment. Financial and infrastructure constraints also feature in pathway A. Unequal adaptive capacity (pathway D) captured differences in autonomy, mobility, role and access to resources. A staff account contrasted mobile workers’ difficulty accessing water with ward-based nurses’ difficulty leaving clinical areas. Another described tension between staff cooling needs and a warm clinical environment. Patient/public analysis extended this to dependency on others and inability to alter clothing, positioning or surroundings. Preparedness and business-continuity underuse (pathway E) combined dispersed ownership with the availability of established planning structures. Institutional framing (pathway F) concerned the potential legitimacy of familiar operational language and trusted clinical voices. National stakeholder accounts were particularly influential in these pathways. Their proposed routes to improved coordination or engagement remain prospective explanations, rather than demonstrated intervention effects.

Together, the six pathways provided an initial explanation of why overheating can persist despite recognition. Pathway A located the problem in a gap between awareness and delivery capacity. Pathway B explained why the evidential signal reaching governance can remain weak. Pathway C showed how essential short-term adaptation can preserve care while leaving structural conditions unchanged. Pathway D established that burden depends on the interaction among exposure, embodied sensitivity, occupational or clinical demand and environmental control. Pathway E identified preparedness and business continuity as available but incompletely activated coordination mechanisms. Pathway F explained how institutional language and trusted voices can affect legitimacy and implementation traction.

### Integrated systems explanation via AcciMap and national stakeholder refinement

The integrated theme and pathway analysis indicated that overall, hospital overheating was not caused by a single issue. Instead, it resulted from different factors interacting across the hospital system. These factors ranged from national policies and funding pressures to building design, day-to-day operational decisions, and the experiences of patients and staff. The factors were summarized in our final AcciMap (**Figure 2**), which was intended to identify how different parts of the hospital system interact to create the conditions in which overheating occurs and affects people.

**Figure 2.**
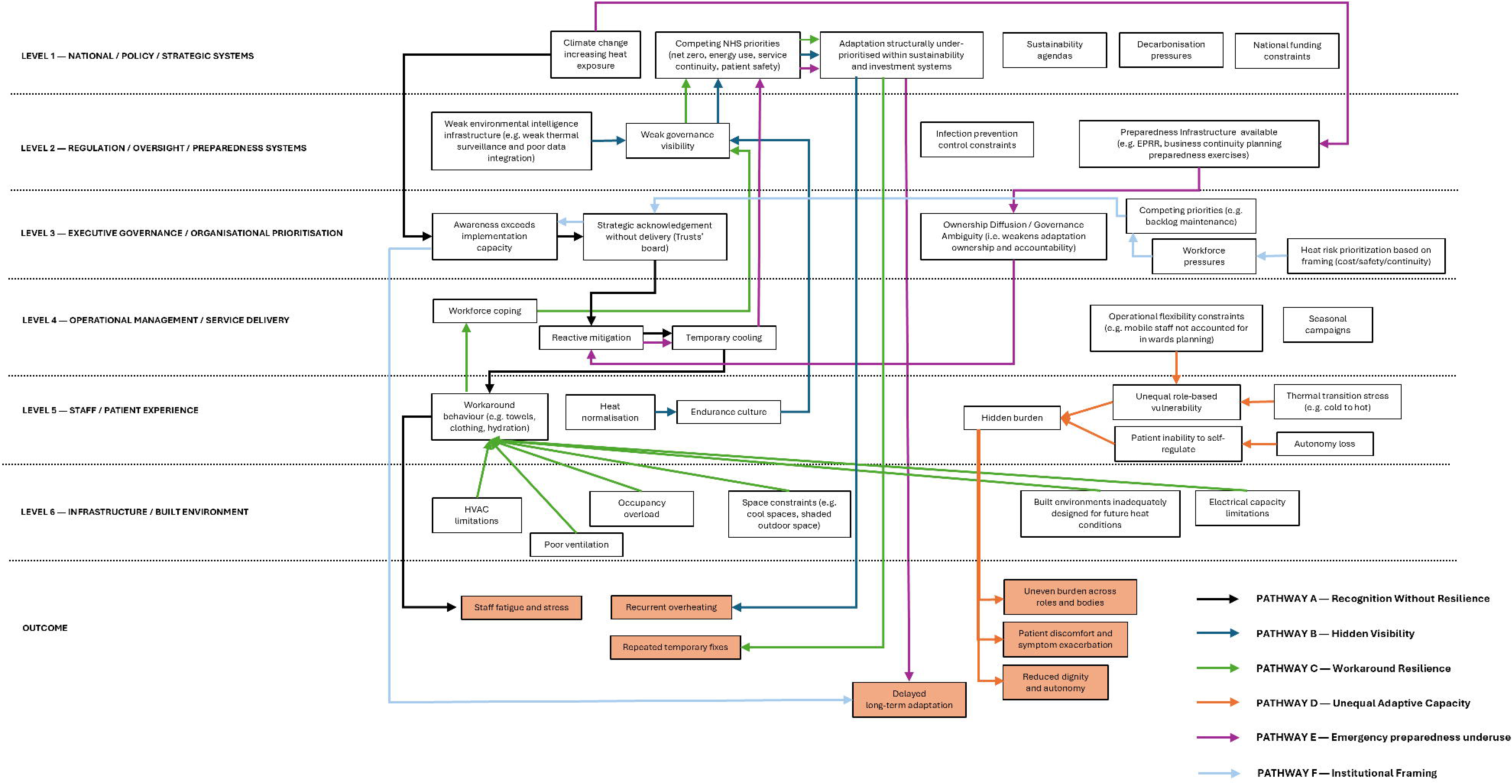
AcciMap of hospital overheating risk and unequal thermal burden. Factors are organised across six interacting system levels and an outcome band. Coloured lines correspond to the six pathways. Factors may contribute to more than one pathway; arrows represent evidence-supported explanatory relationships rather than statistically tested causal effects.

The map was organised into seven levels that identified where different influences and decisions occur. Each box within the map represents a factor identified through our data analysis across Phases 1 -4. Some factors related to policies, funding and organisational priorities. Others related to buildings, equipment, workplace cultures, patient experiences or operational practices. As noted, no single factor explained overheating on its own. One of the most important features of the map is that factors at different levels could influence one another. For example:

- National priorities can shape organisational decisions.
- Organisational decisions can influence operational practices.
- Operational practices can affect the experiences of patients and staff.
- Experiences of patients and staff can highlight problems that may otherwise remain hidden.

This means that changes in one part of the system can sometimes create benefits elsewhere. Several factors shown on the map were interconnected, meaning that no single organisation or group can solve the problem alone. At the bottom of the map are the outcomes identified through the qualitative data analysis. The coloured lines highlight the six recurring pathways identified via the thematic synthesis, namely:

• Pathway A – Recognition Without Durable Resilience

• Pathway B – Hidden Visibility

• Pathway C – Workaround-based Resilience

• Pathway D – Unequal Adaptive Capacity

• Pathway E – Emergency Preparedness and Business Continuity Underuse

• Pathway F – Institutional Framing

It is important to note that these pathways are not the only way to read the map. Instead, they help focus on different ways that overheating develops and persists within the hospital settings studied. Furthermore, the pathways should not be viewed in isolation. Many factors appear in more than one pathway, and many pathways contribute to the same outcomes. For example:

• Better monitoring could improve visibility of overheating while also helping organisations take earlier action.

• Preparedness planning could strengthen resilience across multiple pathways.

• Supporting staff and patients may reduce unequal impacts while also improving organisational awareness of overheating risks.

Because the pathways interact, considerations around potential interventions should consider how intervening in one area may create benefits elsewhere in the system. As such, pathways can be viewed as “entry points” for meaningful actions, interventions, and change.

The final national stakeholder focus group confirmed that the AcciMap was structurally stable, theoretically mature, and highly coherent. Furthermore, two documented refinements show how national stakeholder input changed some explanations. First, ownership diffusion sharpened pathway E by explaining why responsibility spread across disciplines could leave responses reactive. Second, trusted clinical voices and the language of “seasonal pressures” sharpened pathway F as candidate means of improving organisational engagement. These refinements retained the broad pathway structure while changing its explanatory specificity.

The AcciMap enabled visualisation of the system’s complexity and suggested that hospital overheating persists because material vulnerability, fragmented governance, weak intelligence, competing priorities and unequal adaptive capacity interact; continuity is maintained through local coping that can conceal burden and defer durable adaptation; and resilience requires a shift towards anticipatory, monitored and equitable action across governance, preparedness, service delivery and infrastructure.

## Discussion

### Principal findings

This study reframes hospital overheating in England as a patient-safety and socio-technical adaptation challenge. Its central finding is not that hospitals or staff fail to recognise heat risk. Rather, our analysis indicated that recognition can be disconnected from the authority, evidence, resources and coordination required for durable action. The six pathways explained how that gap is reproduced and how its consequences are distributed unequally within the settings investigated.

Climate-resilience frameworks identify buildings, monitoring, workforce capability, governance and preparedness as necessary components of a well-adapted health system.^[5,10,11,16,17,29,32]^ The study by Brooks and colleagues similarly identified disrupted work, competing priorities and preparedness limitations during hot weather in English hospitals.^[18]^ Our findings extend this literature by explaining how these components interact: policy recognition and local awareness can coexist with fragmented authority, limited information and recurrent reliance on informal adaptation. Our contribution is therefore not an inventory of barriers, but an empirically traceable account of the mechanisms through which acknowledged heat risk remains operationally persistent.

### How the pathways combine

Our analysis indicated that recognition without durable resilience is generated across levels. Hospital estates teams cannot independently resolve capital constraints; frontline teams cannot redesign buildings; sustainability leads may not control operational preparedness; and executive concern is weakened when reliable evidence of indoor exposure and consequence is unavailable. This distributed constraint explains persistence more convincingly than attributing failure to organisational indifference or a single department. It is consistent with socio-technical accounts in which safety outcomes arise through interactions among system components rather than a proximal failure,^[19,21,35]^ and extends previous qualitative evidence of competing priorities in hospitals during hot weather by identifying a recurrent mechanism linking short-term operational pressure with long-term under-adaptation.^[18]^

Hidden visibility and workaround-based resilience form a reinforcing relationship. Resilient-healthcare scholarship emphasises the adjustments through which staff sustain care under variable conditions and distinguishes Work as Done from formal Work as Imagined.^[23,25]^ Our data demonstrate an important qualification: the same adjustments can transfer adaptation costs to individuals and reduce the signals reaching decision-makers. A fan may offer relief while creating space or infection-control difficulties; staff may alter workflow or tolerate strain without recording it; and successful coping may prevent an observable service failure. Fewer incidents may therefore reflect resignation, normalisation or invisible work rather than lower burden. This finding does not characterise workarounds as intrinsically harmful. It identifies the conditions under which necessary adaptation ceases to generate organisational learning.

The patient-safety implication is that continued service cannot be treated as a sufficient indicator of resilience. Heat-related fatigue and reduced work capacity have plausible implications for performance,^[12–15]^ but conventional incident systems may capture only overt failures. Assessment should also document symptoms, repeated informal adjustments, inaccessible mitigation, patient discomfort and loss of autonomy. The relevant question is not only whether care stopped, but what staff and patients had to do—and what burden they absorbed—to prevent it from stopping.

Unequal adaptive capacity adds an equity dimension to this dynamic. Our data indicated that vulnerability was not located solely in an individual’s diagnosis or in room temperature. It arose through the relationship among exposure, physiology, occupational or clinical demand and capacity to alter the environment. This interpretation is compatible with climate-risk frameworks that distinguish hazard, exposure and adaptive capacity,^[5,29]^ and with evidence that thermal sensitivity varies across bodies and life stages.^[31]^ The study adds hospital-specific mechanisms: transient work, safe-staffing constraints, patient dependency, restricted movement and loss of control over clothing, bedding, airflow or location.

Aggregate continuity and mean temperature can therefore conceal who cannot leave, hydrate, change clothing, access recovery space or request assistance. Workforce adaptation literature highlights preparedness, competencies, leadership and multisectoral planning, ^[30]^ but nominal provision does not establish equitable access. A hydration point or cooling space may exist while remaining inaccessible to a worker moving across wards or unable to leave a clinical area. Monitoring should ask who is exposed, who can act, who requires assistance, who absorbs the cost of continuity and whose experience enters formal data.

The pathways E and F identified routes to implementation. The Adverse Weather and Health Plan for England supplies a national alert architecture,^[17]^ and NHS adaptation and wider climate-resilience frameworks call for preparedness, workforce capability and resilient facilities.^[10,11,16,29,32]^ Our evidence indicates why these resources may remain underused unless heat is connected to locally explicit triggers, roles, action cards, exercises and review. Preparedness can create a learning loop between forecast, operational action, post-event evidence and structural escalation.

Framing matters because “comfort” can imply discretion, while safety, workforce health, seasonal pressure and continuity connect overheating to established governance duties. ^[22]^ The stakeholder evidence supports this as a plausible implementation mechanism, not a tested communication intervention. Reframing should not displace investment in buildings or dilute the specificity of increasing heat risk. Its potential value lies in creating institutional legitimacy for coordinating immediate protection, seasonal preparedness and long-term estate adaptation.

These relationships also explain why the six pathways should not be interpreted as six parallel problems. Recognition can remain organisationally weak when hidden visibility limits evidence and institutional framing limits legitimacy. Workarounds can compensate for insufficient preparedness while simultaneously concealing unequal burden. Conversely, embedding heat within preparedness can improve visibility by defining what should be monitored, who should receive the information and what response should follow. Our AcciMap therefore represents feedback and dependency: interventions directed at a shared node may influence several pathways, while action on one pathway alone may be neutralised by constraints elsewhere.

The interaction between time horizons is equally important, particularly to inform future iterations of the Adverse Weather and Health Plan for England. Immediate protective action cannot await estate renewal, but repeated short-term coping should generate evidence and escalation for seasonal and structural change. Seasonal planning can connect forecasts and alerts to staffing, cooling access and surveillance, while post-event review can identify where infrastructure investment is required. Without this learning loop, an apparently effective immediate response may reproduce the conditions that make the same response necessary the following year.

### Implications for improvement

Our AcciMap indicated that isolated interventions are unlikely to be sufficient. Systems-thinking guidance and AcciMap research emphasise that interventions entering one part of a complex system can be enabled or neutralised by conditions elsewhere.^[20,33,48]^ Durable adaptation therefore requires a portfolio operating across time horizons. Immediate measures include accessible hydration, cooling and staffing adjustments; seasonal measures include monitoring, heat plans, training, business -continuity exercises and predefined escalation; structural measures include ventilation, estate redesign, electrical capacity and capital investment. These are linked functions rather than interchangeable solutions.

Environmental intelligence is a key entry point, but sensors alone will not produce learning. National and international guidance supports monitoring and vulnerability assessment, ^[9,11,29,32]^ yet data require thresholds, ownership and a defined response. Indoor exposure should be connected with staff symptoms, patient experience, workarounds, incidents, service effects and accountable decisions. Technical and qualitative evidence are complementary: monitoring establishes patterns, while staff and patient accounts reveal control, dependency and operational consequence. Evaluation should assess whether action reduces exposure and displaced burden, not merely whether services continue.

Co-production is similarly more than consultation. Staff and patients possess situated knowledge of thermal transitions, clinical trade-offs and inaccessible mitigation. Their participation can identify why a nominally available intervention fails for particular roles or conditions. Intervention design should specify the intended function, locally appropriate delivery form, ownership, dependencies, equity implications and hypothesised CMO logic before prospective testing. Realist-informed evaluation would then ask whether the expected mechanism was activated in the local context, rather than judging only whether the intervention was present.^[24,28]^

### Methodological contribution and transferability

AcciMap was developed for analysing accidents across complex socio-technical systems.^[20,33]^ We extended it to recurrent climate vulnerability by integrating operational reports, organisational perspectives and lived experience with realist-informed explanation. The six pathways prevent the map from becoming a static inventory: they show how vulnerability is reproduced and how shared nodes may serve as intervention entry points. This use is consistent with systems approaches to health-system strengthening^[34]^ and with healthcare research calling for analysis beyond proximal individual actions.^[21,35,36]^

Our current approach could inform a future practical mapping tool and investigation in other sectors where environmental hazards interact with service obligations and unequal autonomy (e.g. consider the educational and social care sectors). However, such applications require local evidence and evaluation of usability and transferability. For example, our AcciMap should not be transferred unchanged. Hospitals differ in estates, services, climate exposure, governance and adaptive resources. Transferability lies in the process of local remapping and stakeholder challenge, not in assuming universal factors or links.

AcciMap also does not quantify exposure, estimate prevalence or establish intervention effectiveness. It should connect, rather than replace, building-performance analysis, epidemiology, physiology, economics and prospective implementation research. The six pathways are therefore sensitising propositions for local testing, not a universal causal model.

### Strengths and limitations

Triangulation across 129 operational reports, formal plans, interviews, staff and patient focus groups and national stakeholders connected Work as Imagined with Work as Done. The displayed cross-source matrix, CMO-informed pathway formulations, quotations and successive maps make the evidential chain inspectable. Combining systems mapping with realist-informed reasoning moved the analysis from a list of barriers to an explanation of recurrent outcomes.

The primary empirical sites were two hospital trusts in one English region. National stakeholders strengthened the relevance of governance and implementation findings but did not validate every factor or link across the NHS. Incident data were created for operational purposes, differed by reporting system and period, and were affected by the under-reporting identified in the analysis. Purposive qualitative sampling captured cross-level perspectives but did not represent all staff groups, patients or decision-makers; nor our data sources reflected all possible outcomes likely to arise from heat stress (e.g. increased patients’ mortality within hospitals during heatwaves). Finally, the AcciMap contains evidence-supported explanatory propositions, not statistically tested causal relations. Prospective work should examine the model across diverse hospitals and test whether mechanism-targeted interventions change exposure, burden, equity and service outcomes.

## Conclusion

Our analysis indicated that overheating in hospitals in England persists not because the risk is unknown, but because capacity to act is fragmented across infrastructure, governance, information, preparedness and everyday service delivery. Workarounds maintain care but can conceal the conditions under which continuity is achieved, while differences in role, physiology, mobility and autonomy distribute that burden unequally. Durable resilience therefore requires a shift from recurrent coping towards anticipatory, monitored and equitable adaptation. Our AcciMap supports that shift by making cross-level interactions visible and converting them into locally testable intervention propositions. Their wider value is not a universal map of hospital overheating, but a transferable process through which healthcare organisations can diagnose climate vulnerability, challenge assumptions and design action proportionate to system complexity.

## Data Availability

All data produced in the present study are available upon reasonable request to the authors

## Acknowledgements

We wish to acknowledge all participants in the research for volunteering their time and perspectives.

